# Prediction of *TP53* biomarkers and survival outcomes from whole slide images using a vision transformer-based multi-instance learning framework

**DOI:** 10.1101/2025.11.11.25340052

**Authors:** Abadh K Chaurasia, Patrick W Toohey, Matthew T Bennett, Helen C Harris, Alex W Hewitt

**Affiliations:** Menzies Institute for Medical Research, University of Tasmania, Australia; Pandani Solutions Pty Ltd, Hobart, Australia; Department of Pathology, Royal Hobart Hospital, Hobart, Australia; School of Medicine, University of Tasmania, Australia

**Keywords:** Computational Pathology, Multiple-instance learning, Pan-cancer, Precision Oncology, Survival Analysis, *TP53* mutation, Vision Transformer, Whole Slide Images

## Abstract

**Background:** Accurate molecular profiling and prognostication from routine histopathology slides could transform precision oncology. We developed a Vision Transformer (ViT)-based multi-instance learning (MIL) framework for combined predictions of 32 solid tumour types, *TP53* biomarker detection, and survival prediction directly from Whole Slide Images (WSIs).

**Methods:** 11,060 primary tumours were curated from the TCGA Pan-Cancer Atlas with corresponding somatic mutations, RNA-seq, and clinical outcome data. *TP53* alterations were classified as pathogenic drivers using COSMIC and hotspot annotations. WSIs underwent tissue masking, quality control, stain normalisation, and patch extraction (518 x 518) at 6x downsampling. Each patch was encoded by a ViT into a 768-dimensional embedding, which formed a token sequence for a 6-layer Transformer aggregator with learnable classification and positional embeddings. Seven task heads were developed to generate predictions for various outcomes, including cancer type, *TP53* mutation status, *TP53* RNA expression levels, overall survival (OS), progression-free interval (PFI), and the corresponding times for OS and PFI. The training process had two stages. First, the model was trained on tumour tissue patches from WSIs at five magnifications. In the second stage, it was fine-tuned using patches from all tissue regions with a content-aware strategy, updating all MIL layers for a maximum of 150 epochs at a learning rate of 1 × 10⁻⁵. The model’s performance was evaluated on an independent validation set of 1,729 slides using classification metrics, including the area under the receiver operating characteristic curve (AUROC), regression metrics, and Concordance indices (C-index).

**Results:** The multi-resolution ViT-based MIL model achieved an AUROC of 0.775 (95% CI: 0.749–0.801) for *TP53* mutation detection on the validation set, demonstrating strong overall performance across classification and survival prediction tasks. The fine-tuned model attained robust performance across the tasks, with 0.7569 accuracy for cancer classification, 0.745 AUROC for *TP53* mutation detection, C-indices of 0.686 and 0.650 for OS and PFI, and a mean squared error of 1.072 for *TP53* RNA expression level estimation. The fine-tuned model attained an accuracy of 65.9% (95% CI: 0.636–0.681) in tumour classification and an AUROC of 0.766 (95% CI: 0.743–0.789) for detecting *TP53* mutations on the external validation set. However, most tumour classes, aside from ovarian cancer, reached an AUROC above 0.88 with class-specific thresholding using the Youden Index. This indicates strong generalisation across 32 tumour types, providing reasonable molecular profiling but offering limited prognostic utility in surgical oncology.

**Conclusion:** A ViT-based MIL model can simultaneously infer tumour taxonomy, *TP53* mutation status, and *TP53* RNA expression levels directly from WSIs, with performance comparable to conventional genomic assays, while prognostic risk remains limited. This integrated, slide-level approach offers a scalable pipeline toward computational pathology.

## INTRODUCTION

Histopathology remains the gold standard for diagnosing cancer and identifying prognostic features across most solid tumours.^1,2^ While routine Haematoxylin and Eosin (H&E) stained Whole Slide Images (WSIs) capture the comprehensive morphological spectrum of tumour architecture, current clinical workflows still depend on additional molecular and genomic assays to identify key alterations such as *TP53* mutations.^3–6^ *TP53*, the most commonly altered tumour suppressor gene across human cancers, is an influential genomic biomarker.^7–9^ Pathogenic driver mutations in *TP53* disrupt genomic stability, cell-cycle regulation, and apoptosis, influencing both tumour behaviour and treatment resistance.^10^ Thus, accurate diagnosis and molecular profiling are essential for guiding treatment and improving patient outcomes. Additionally, survival outcomes, including overall survival (OS) and progression-free interval (PFI), are equally important in guiding therapeutic decisions.^11^ However, these require separate assays: histopathology for tumour classification, sequencing for mutation detection, and statistical modelling for survival analysis — each of which is resource-intensive and often inaccessible in low-resource settings, leading to diagnostic delays and inequitable access to precision oncology.

Recent advances in computational pathology and deep learning have enabled the extraction of clinically relevant morphological, prognostic, and molecular features directly from WSIs.^12^ These technologies demonstrate promising results in automating tumour classification, biomarker prediction, and survival analysis. However, most existing approaches emphasise single-task objectives and often remain limited to specific cancer types, thus failing to capture the broader potential of integrative pan-cancer analysis.^13–16^ Moreover, these models typically require large, well-annotated datasets, which are difficult to obtain in histopathology due to the high volume of WSIs and the lack of detailed region-level annotations. This presents a significant challenge for traditional supervised learning methods, which rely on precise annotations to guide model training. In this context, a multi-instance learning (MIL) framework for computational pathology allows models to learn from slide-level labels by aggregating information from multiple patches within each slide, without the need for thorough manual annotation.^17^

MIL frameworks demonstrate strong performance across various slide-level tasks, including cancer subtype classification, mutation prediction, and survival modeling.^18,19^ Among these, attention-based MIL models are particularly effective at capturing both local and global morphological patterns, enabling nuanced interpretation of tumour heterogeneity.^20^ In parallel, self-supervised learning methods learn general-purpose visual representations from large-scale unlabeled datasets, which can be fine-tuned for downstream pathology tasks.^21–23^ To our knowledge, a comprehensive Vision Transformer (ViT)-based MIL framework that simultaneously addresses tumour classification, mutation prediction, *TP53* RNA expression quantification, and overall survival modelling has not yet been developed. This leaves a critical gap in the translation of multitask MIL frameworks into a unified, clinically relevant tool that combines diagnostic, molecular, and prognostic information within computational pathology.

Herein, we present a multitask, ViT-based MIL framework capable of jointly predicting seven clinically relevant parameters from H&E-stained WSIs. These include pan-cancer classification, *TP53* mutation status, *TP53 RNA* expression level, and four survival-related outcomes. Our approach integrates diagnostic, molecular, and prognostic tasks, associating morphological evaluation with genomic analysis to advance morpho-genomic modelling and support precision oncology across diverse solid tumours.

## METHODS

This study presents a transformer-based MIL model for extracting diagnostic, molecular, and prognostic insights from H&E-stained WSIs, as depicted in **Figure 1**.

**Figure 1:**
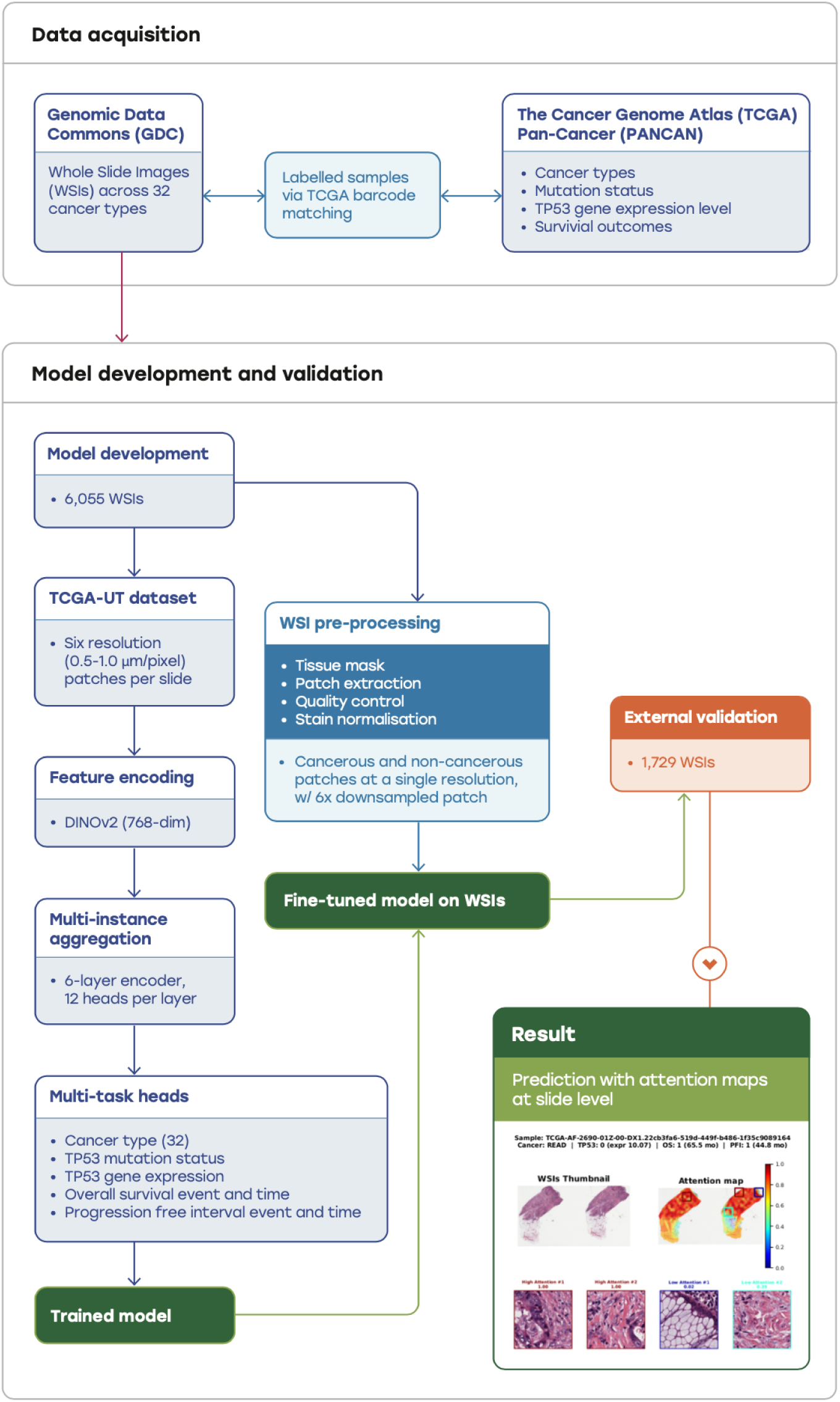
Overview of the study: schematic workflow for developing and validating a multi-task learning model using Whole Slide Images (WSIs).

### Cohorts and datasets

#### TCGA Pan-Cancer (PANCAN)

We extracted a public somatic mutation dataset from the TCGA Pan-Cancer Atlas.^24,25^ It contains ∼3 million mutations across 9,107 tumour samples that were annotated with either pathogenic or wild-type *TP53* status, based on the presence or absence of alterations in the *TP53* gene. *TP53* RNA expression levels were obtained from the batch-corrected RNA-Seq dataset.^26^ The expression values were extracted for each sample and matched to mutation data using TCGA sample barcodes. This dataset contains normalised mRNA expression profiles for 20,532 genes across 11,060 primary tumour samples from 33 cancer types. Expression values were generated using the RSEM pipeline, batch-effect corrected, and reported as log_2_(norm_value+1). Clinical annotations were obtained for 12,591 tumour samples from the Pan-Cancer Atlas.^27^ From this dataset, we extracted cancer type abbreviations and Survival endpoints, including OS and PFI, with corresponding event indicators and follow-up times. Finally, each sample was labelled based on matching barcodes with cancer types, *TP53* genomic variants, gene expression, and survival outcomes.

### *TP53* Mutation Classification

*TP53* mutations were systematically classified into four hierarchical categories for all the slides: pathogenic driver, pathogenic passenger, benign, and wild-type *TP53* mutations **(Supplemental Figure S1)**. A gold-standard driver set was first constructed as the union of all amino acid changes labelled as “Confirmed somatic variant” in the COSMIC Mutant Census (v102) and all variants annotated as “Hotspot = Yes” in the TP53 database.^28,29^ Variants present in this combined set were assigned as Pathogenic drivers. Pathogenic passenger mutations included variants not in the driver set but predicted to be deleterious or damaging by functional consequence (e.g., nonsense, frameshift, splice-site) or in-silico algorithms (deleterious by Sorting Intolerant From Tolerant (SIFT) or possibly/probably damaging by Polymorphism Phenotyping (PolyPhen). Variants with no evidence of pathogenicity, including synonymous changes, noncoding alterations, or variants predicted as benign by both SIFT and PolyPhen, were assigned to the Benign class. Finally, samples with no *TP53* mutation reported in the TCGA MAF file were grouped with the benign variants. At the sample level, a hierarchical rule was applied such that the most severe mutation present determined the final label (Pathogenic driver > Pathogenic passenger > Benign > No *TP53* Mutation). Mutation processing, annotation, and classification were performed using custom Python scripts.

### TCGA-UT Dataset

This dataset contains over 1.6 million histological cancerous patches only, extracted from 8,736 diagnostic H&E-stained WSIs from 7,175 patients across 32 solid tumour types.^30^ The patches were taken from histologically uniform tumour regions, confirmed by two pathologists, and low-quality slides were excluded during quality control. From each annotated region, 10 patches were randomly sampled at six magnification levels (0.5-1.0 μm/pixel), providing a multi-resolution representation of tissue morphology (**Supplementary Figure S2**). This multi-resolution dataset captures morphological details at various tissue resolutions, making it a valuable resource for computational pathology.

### The Cancer Genome Atlas (TCGA) dataset

WSIs were downloaded from the Genomic Data Commons (GDC) Data Portal for the sample involved in PANCAN to capture their natural histopathology morphology, including cancerous and non-cancerous tissues. The selection was restricted to cases across 32 solid tumour types for which corresponding molecular and clinical annotations (mutation status, gene expression, and survival outcomes) were available in the TCGA Pan-Cancer Atlas. By aligning to the curated sample set, only WSIs from patients with complete *TP53* mutation profiles, RNA-seq expression data, and harmonised clinical endpoints were included in this study, resulting in 7,784 WSIs. These WSIs were then subjected to patch extraction, stain normalisation, and ViT-based feature encoding, providing standardised inputs for our MIL framework.

### WSIs preprocessing

The preprocessing pipeline for WSIs was designed to detect the tissue within a slide, extract patches, and implement strict quality control at the patch level before feature encoding. Each WSI was processed at a low-resolution pyramid level to construct a binary tissue mask. The downsampled RGB image was converted to grayscale and thresholded, with darker pixels considered tissue and lighter pixels considered background. Morphological closing, opening, and dilation operations were applied to smooth contours and fill small gaps. Small connected components (area <5,000 pixels at the mask level) were discarded to remove debris and spurious detections. The mask level and native level-0 geometry were recorded to allow coordinate mapping during tiling. This mask was then upscaled to the corresponding coordinates at the native slide resolution and used to guide subsequent patch selection. Slides were then traversed on a non-overlapping grid whose stride matched the intended field of view. Patches were extracted at an effective 6x downsampling relative to the native resolution, with a fixed size of 518 × 518 pixels. Further, the patches were required to contain sufficient tissue (≥20%) and were rejected if dominated by a near-white background (>90% of pixels near white). Sharpness was quantified by the variance of the Laplacian, and patches that were blurred or out of focus (variance < 100) were discarded. To suppress ink and dark artifacts, patches were transformed to HSV space and rejected if the fraction of very low-saturation, low-value pixels exceeded 2%. Only patches meeting all the criteria were retained, resulting in per-slide collections of artifact-free, tissue-rich patches downsampled to 518 x 518 pixels.

### MIL architecture and Transformer encoder

The weakly supervised MIL architecture utilised a frozen self-supervised ViT backbone (DINOv2; 768-dimensional embeddings) for patch encoding, paired with a six-layer Transformer encoder (12 self-attention heads per layer, feed-forward width 4d, and dropout 0.1) for slide-level aggregation.^31,32^ For each slide, patch embeddings were arranged into a token sequence with a learnable classification (CLS) token prepended and positional embeddings added; the final hidden state of the CLS token was used as the slide-level representation. Task-specific heads formed even outputs: cancer type, *TP53* mutation, *TP53* RNA expression levels, OS event, PFI event, and two time-to-event (OS time and PFI time).

### Training Strategies

Slide-level training was conducted in two stages, using the same set of TCGA data and patient-level split (80% training, 20% validation) in both stages, but differing in whether slides contained non-cancerous tissues.

### Stage 1: Multi-resolution ViT-based MIL model

In this stage, the model was trained only on the tumour-containing tissue across six magnifications (0.5–1.0 μm/pixel) of WSIs from the TCGA-UT Dataset. Reinhard stain normalisation was applied to each slide using a fixed reference, followed by light augmentation (resize, horizontal/vertical flips, mild colour jitter) for most classes. To address class imbalance, minority cancer types acquired aggressive augmentation, including additional rotations, affine transformations, and increased colour jitter. For each slide, a fixed-length bag of 300 tokens was constructed by randomly subsampling embeddings when more were available or by padding and masking when fewer were present. We initially trained only the MIL heads with the aggregator frozen for up to 100 epochs using a learning rate (LR) of 2 × 10⁻³. This was followed by fine-tuning all MIL layers for another 100 epochs at a reduced LR of 2 × 10⁻⁵. Throughout both phases, we applied a weight decay of 2 × 10⁻⁴ and used a cosine LR schedule with a 5% warm-up, gradually increasing the LR from 0 to its peak before decaying to 2 × 10⁻⁷. Mini-batches were constructed using a BalancedBatchSampler^33^ to ensure an approximately uniform mix of cancer types per batch. Additionally, class weights for the cancer head were recomputed at each epoch, scaled by a factor of three to address class imbalance.

### Stage 2: Fine-tuning model on WSIs (tumour + non-tumour tissue)

At this stage, the Multi-resolution ViT-based MIL model was fine-tuned on the same subset of the training set (4,843), with a particular focus on tissue-containing regions of WSIs, which included both cancerous and non-cancerous areas. All patches were extracted at a single resolution, approximately corresponding to a 6x downsample. The spatial arrangement of patches from a single WSIs is shown in **Supplementary Figure S3**. The patches were normalised and augmented as in Stage 1, with stronger transforms applied only to minority classes.

During training, each WSIs bag consisted of 300 tokens. For validation, we used a deterministic Top-K selection to ensure stable and reproducible evaluation.^34^ For training, we adopted a hybrid strategy: approximately 70% of the patches were selected based on the highest similarity scores (Top-K), while the remaining 30% were randomly sampled from the remaining patches from the WSIs. This approach promotes exploration and helps prevent overfitting to a fixed subset of instances. The fine-tuned model updated all MIL layers for up to 150 epochs (AdamW, LR = 1×10⁻⁵), with batch size = 64, gradient accumulation (x2), cosine LR schedule (5% warm-up; min LR = 2×10⁻⁷), weight decay 2 x 10⁻⁴, and early stopping (patience = 10). We maintained an exponential moving average (EMA) of model weights with a decay of 0.999 and selected the best EMA model on the validation set for reporting.

### Loss functions

The model was trained using a multi-task objective on slide-level embeddings. For cancer type classification, focal cross-entropy (gamma = 2.0) was applied with label smoothing (epsilon = 0.02) and epoch-wise inverse-frequency class weights (3x). The binary heads for *TP53* mutation and OS/PFI events utilised focal binary cross-entropy (BCE) with logits (gamma = 1.5). Time-to-event outcomes were modelled with Cox partial likelihood, while continuous heads for *TP53* expression and OS/PFI time used Smooth L1 loss. The total loss was a weighted sum of the cancer, binary, Cox, expression, and time terms.

### Slide level Survival analysis

OS and PFI were evaluated based on slide-level prediction outputs, with time measured in months. For each endpoint, inclusion in the analysis required a minimum of two samples per cancer type and at least one observed event; cohorts failing to meet these criteria were excluded. For each cancer type, samples were categorised into risk groups based on a median split of the model’s predicted time-to-event. Samples with a predicted time less than or equal to the cohort median were labelled as High Risk, while those with a predicted time greater than the median were labelled as Low Risk. We estimated Kaplan-Meier (KM) curves for the two groups and compared them using a two-sided log-rank test.^35^

Quantifying the effect size involved fitting a Cox proportional hazards model with a single binary covariate (High vs. Low Risk), reporting the hazard ratio (HR) along with the 95% confidence interval (CI). To enhance stability in small or nearly separated cohorts, a penalised Cox model was employed (with a penalizer of 0.1) and robust variance was used. The model’s discrimination was summarised using Harrell’s concordance index (C-index)^36^, computed on the same cohort.

### Slide-level predictions and interpretability analyses

Predictions were made at the slide level using the trained fine-tuned model, and attention visualisations were generated directly from WSIs. The WSIs’ preprocessing, stain normalisation, and transformation steps were aligned with the previously outlined, which included tissue detection, patch extraction, and quality control. The model generated seven outputs: cancer types (32), *TP53* mutation status (*TP53* pathogenic or *TP53* wildtype), *TP53* RNA expression levels, OS, PFI, and prediction time for both OS and PFI.

To visualise model interpretability, each patch was scored using attention rollout from the MIL transformer.^37^ Attention heads were averaged across layers, identity connections added, rows normalised, and matrices multiplied. The resulting attention flow from the [CLS] token to each patch yielded per-patch importance scores. Patch scores were mapped to slide coordinates, rescaled to a 0–1 range using the 2nd–98th percentiles, and interpolated over a downsampled WSI thumbnail. Gaussian smoothing and tissue masking refined the resulting map. Final heatmaps were overlaid using a turbo colourmap, with the two highest and two lowest attention patches highlighted for visual comparison.

In our study, at the cohort level, each slide was represented by a single feature vector derived from the fine-tuned MIL model’s [CLS] embedding. These vectors were embedded into two dimensions using t-distributed Stochastic Neighbour Embedding (t-SNE)^38^, enabling visual inspection of slide-level relationships across the pan-cancer dataset.

### Statistical analysis and model evaluation

Model development and training were performed using a single NVIDIA GPU with 80 GB of VRAM through a Python script utilising the PyTorch framework.^39,40^ All statistical analyses were conducted at the slide level, using 95% bootstrap CI estimated from 2,000 resamples.^41^ For multiclass cancer-type prediction, the standard approach involved assigning each slide to the class with the highest softmax probability using the argmax function.^42^ To address class imbalance and heterogeneity, a threshold-optimised one-versus-rest (OvR) multiclass approach was applied, with probability thresholds for each cancer type selected to maximise the Youden Index.^43,44^ For binary endpoints, including *TP53* mutations, OS, and PFI events, the area under the receiver operating characteristic curve (AUROC), accuracy, sensitivity, and specificity were reported. For the continuous endpoint of *TP53* expression level, the mean squared error (MSE) was computed using regression metrics.

## RESULTS

### Multi-resolution MIL model performance

The multi-resolution ViT-based MIL model demonstrated strong performance, achieving an overall accuracy of 0.7043 (95% CI: 0.6782–0.7302) in classifying WSIs into 32 solid tumour types on the validation set (1212 WSIs) using argmax predictions. When per-class thresholds were applied based on the Youden Index, single-label classification accuracy improved to 0.7230 (95% CI: 0.6972–0.7483). Although OvR AUROCs were notably higher than 0.87 for all cancer types. All classification metrics are presented in **Supplementary Table S1,** while the confusion matrix for OvR classes is displayed in **Supplementary Figure S4**. For *TP53* mutation detection, overall performance at the Youden Index threshold yielded an AUROC of 0.775 (95% CI: 0.749–0.801), an accuracy of 0.706 (95% CI: 0.681–0.732), sensitivity of 78.6% (95% CI: 74.8–82.4), and specificity of 66.3% (95% CI: 63.0–69.6). Survival discrimination from risk heads showed a C-index of 0.642 (95% CI: 0.608–0.675) for OS and 0.634 (95% CI: 0.605–0.663) for the PFI. *TP53* RNA expression levels attained MSE of 0.944 (95% CI: 0.853–1.041).

### Fine-tuning model performance

The fine-tuned model, trained on both tumour and non-tumour tissue regions of the WSIs, demonstrated strong generalisation across the validation set of 1212 WSIs. Using argmax predictions, it achieved an overall accuracy of 0.7550 (95% CI: 0.7303–0.7783). Performance improved further with class-specific thresholding using the Youden Index, achieving an accuracy of 0.7569 (95% CI: 0.7328–0.7808). The OvR AUROC, accuracy, sensitivity, and specificity scores remained consistently high across all cancer types (**Table 1**). The model for detecting *TP53* mutations achieved an AUROC of 0.745 (95% CI: 0.718–0.771), accuracy of 0.648 (95% CI: 0.621–0.675), sensitivity of 83.3% (95% CI: 79.7–87.0), and specificity of 54.8% (95% CI: 51.2–58.3). The confusion matrix cross-tabulation is shown in **Supplementary Figure S5**, displaying the counts of true *TP53* wildtype and *TP53* pathogenic variant cases versus the predicted wildtype or pathogenic variant statuses for each cancer type. Prognostic risk yielded a C-index of 0.686 (95% CI: 0.654–0.718) for OS and 0.650 (95% CI: 0.623–0.678) for PFI. Additionally, analysing *TP53* RNA expression levels resulted in an MSE of 1.072 (95% CI: 0.938–1.205). These results highlight the model’s strong performance in classification, mutation detection, and prognostic tasks, emphasising its potential utility in computational pathology workflows.

**Table 1.** Fine-tuned model’s performance on the validation set using Youden Index with class-specific thresholds.

| Cancer Class | Slides number | AUROC | Accuracy | Sensitivity | Specificity |
| --- | --- | --- | --- | --- | --- |
| ACC | 10 | 0.987 (0.974–0.997) | 0.962 (0.951–0.973) | 1.000 (1.000–1.000) | 0.961 (0.950–0.972) |
| BLCA | 57 | 0.985 (0.976–0.992) | 0.944 (0.931–0.957) | 0.947 (0.883–1.000) | 0.944 (0.931–0.958) |
| BRCA | 114 | 0.98 (0.965–0.992) | 0.939 (0.926–0.951) | 0.939 (0.891–0.980) | 0.939 (0.925–0.953) |
| CESC | 37 | 0.961 (0.930–0.986) | 0.933 (0.918–0.947) | 0.921 (0.828–1.000) | 0.934 (0.920–0.947) |
| CHOL | 6 | 0.887 (0.655–0.999) | 0.99 (0.984–0.995) | 0.834 (0.429–1.000) | 0.991 (0.985–0.996) |
| COAD | 39 | 0.993 (0.988–0.996) | 0.96 (0.949–0.970) | 1.000 (1.000–1.000) | 0.958 (0.947–0.969) |
| DLBC | 4 | 0.999 (0.997–1.000) | 0.997 (0.993–0.999) | 0.983 (1.000–1.000) | 0.997 (0.993–0.999) |
| ESCA | 22 | 0.979 (0.963–0.991) | 0.892 (0.874–0.910) | 1.000 (1.000–1.000) | 0.889 (0.871–0.907) |
| GBM | 17 | 0.996 (0.992–0.999) | 0.977 (0.968–0.985) | 1.000 (1.000–1.000) | 0.977 (0.967–0.985) |
| HNSC | 59 | 0.984 (0.972–0.994) | 0.956 (0.945–0.967) | 0.949 (0.890–1.000) | 0.956 (0.944–0.967) |
| KICH | 9 | 0.997 (0.994–1.000) | 0.988 (0.982–0.994) | 1.000 (1.000–1.000) | 0.988 (0.982–0.994) |
| KIRC | 53 | 0.995 (0.989–0.999) | 0.973 (0.963–0.981) | 0.962 (0.906–1.000) | 0.973 (0.963–0.982) |
| KIRP | 41 | 0.99 (0.981–0.996) | 0.969 (0.959–0.978) | 0.95 (0.868–1.000) | 0.97 (0.960–0.979) |
| LGG | 92 | 0.998 (0.994–1.000) | 0.979 (0.971–0.987) | 0.978 (0.943–1.000) | 0.979 (0.970–0.987) |
| LIHC | 52 | 0.986 (0.959–0.999) | 0.984 (0.977–0.991) | 0.981 (0.939–1.000) | 0.984 (0.977–0.991) |
| LUAD | 84 | 0.972 (0.953–0.986) | 0.94 (0.926–0.954) | 0.928 (0.867–0.977) | 0.941 (0.927–0.955) |
| LUSC | 84 | 0.962 (0.946–0.975) | 0.867 (0.848–0.886) | 0.917 (0.849–0.973) | 0.863 (0.842–0.882) |
| MESO | 12 | 0.878 (0.724–0.978) | 0.918 (0.902–0.934) | 0.755 (0.462–1.000) | 0.92 (0.905–0.934) |
| OV | 1 | 0.779 (0.758–0.801) | 0.779 (0.756–0.801) | 0.633 (0.000–1.000) | 0.779 (0.756–0.803) |
| PAAD | 23 | 0.973 (0.929–0.997) | 0.95 (0.937–0.961) | 0.957 (0.857–1.000) | 0.949 (0.937–0.961) |
| PCPG | 9 | 0.999 (0.995–1.000) | 0.988 (0.981–0.993) | 1.000 (1.000–1.000) | 0.987 (0.981–0.993) |
| PRAD | 60 | 0.992 (0.977–1.000) | 0.965 (0.955–0.975) | 0.983 (0.943–1.000) | 0.964 (0.953–0.974) |
| READ | 11 | 0.941 (0.905–0.973) | 0.831 (0.810–0.853) | 1.000 (1.000–1.000) | 0.83 (0.809–0.851) |
| SARC | 38 | 0.996 (0.990–0.999) | 0.977 (0.968–0.985) | 0.974 (0.911–1.000) | 0.977 (0.968–0.985) |
| SKCM | 38 | 0.99 (0.983–0.995) | 0.933 (0.919–0.948) | 1.000 (1.000–1.000) | 0.931 (0.916–0.944) |
| STAD | 49 | 0.971 (0.948–0.989) | 0.949 (0.936–0.960) | 0.876 (0.773–0.960) | 0.952 (0.939–0.964) |
| TGCT | 19 | 0.999 (0.997–1.000) | 0.988 (0.982–0.994) | 1.000 (1.000–1.000) | 0.988 (0.982–0.994) |
| THCA | 72 | 0.997 (0.994–0.999) | 0.979 (0.970–0.986) | 0.972 (0.928–1.000) | 0.979 (0.970–0.987) |
| THYM | 15 | 0.996 (0.989–1.000) | 0.954 (0.943–0.965) | 1.000 (1.000–1.000) | 0.954 (0.942–0.966) |
| UCEC | 61 | 0.99 (0.984–0.994) | 0.935 (0.921–0.949) | 1.000 (1.000–1.000) | 0.931 (0.916–0.945) |
| UCS | 10 | 0.962 (0.926–0.991) | 0.862 (0.843–0.882) | 1.000 (1.000–1.000) | 0.862 (0.841–0.881) |
| UVM | 11 | 0.998 (0.994–1.000) | 0.982 (0.974–0.989) | 1.00 (1.000–1.000) | 0.982 (0.973–0.989) |

### External Validation

On an independent validation set (1,729 WSIs across 32 solid tumour types), the fine-tuned model attained an overall accuracy of 0.659 (95% CI: 0.637–0.682) using argmax inference, with comparable performance (0.659; 95% CI: 0.636–0.681) under a Youden-optimised OvR strategy, demonstrating robust generalisation across diverse tissue contexts. Most tumour classes reached an AUROC score above 0.88 with class-specific thresholding using the Youden Index, except for ovarian cancer (OV), as shown in **Table 2**. In the binary task of *TP53* mutation detection, the model executed an AUROC of 0.766 (95% CI: 0.743–0.789), with accuracy of 72.0% (95% CI: 69.8–74.1), sensitivity of 70.0% (95% CI: 66.3–73.9), and specificity of 72.9% (95% CI: 70.3–75.3). The confusion matrix cross-tabulation is shown in **Figure 2**. Prognostic performance dropped substantially on the test set compared to the validation set, with approximately 48% and 41% reductions in OS and PFI discrimination, respectively, indicating limited predictive power for survival-related signals. For *TP53* RNA expression level estimation, the model reached an MSE of 1.213 (95% CI: 1.089–1.343), indicating reasonable alignment between predicted and observed expression levels.

**Figure 2.**
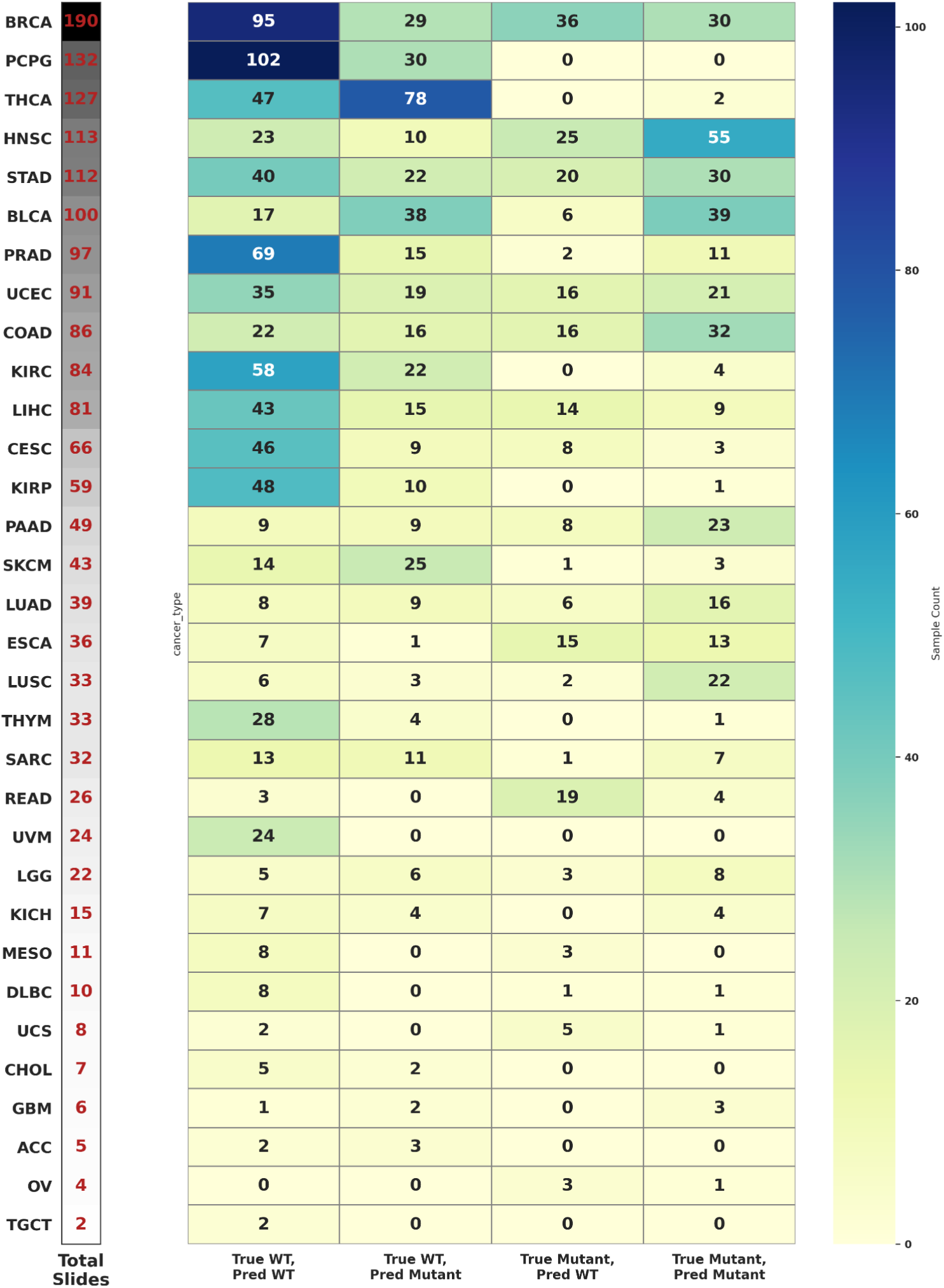
Heatmap of TP53 mutation prediction across pan-cancer on the external validation set. Each row displays a tumour type along with the total number of Whole Slide Images (WSIs), while the columns indicate the true wildtype of TP53 or pathogenic variants (mutant) versus predicted wildtype/pathogenic.

**Table 2.** Performance of the fine-tuned model on the pan-cancer external validation set, estimated classification metrics using class-specific thresholds derived from the Youden Index.

| Cancer Class | Slides number | AUROC | Accuracy | Sensitivity | Specificity |
| --- | --- | --- | --- | --- | --- |
| ACC | 5 | 0.916 (0.780–1.000) | 0.704 (0.681–0.726) | 1 (1.000–1.000) | 0.703 (0.680–0.725) |
| BLCA | 100 | 0.916 (0.876–0.949) | 0.896 (0.881–0.910) | 0.85 (0.777–0.917) | 0.898 (0.883–0.912) |
| BRCA | 190 | 0.979 (0.966–0.989) | 0.941 (0.929–0.951) | 0.932 (0.894–0.965) | 0.942 (0.930–0.953) |
| CESC | 66 | 0.969 (0.956–0.980) | 0.882 (0.866–0.897) | 0.939 (0.877–0.986) | 0.88 (0.864–0.895) |
| CHOL | 7 | 0.922 (0.794–0.994) | 0.912 (0.898–0.925) | 0.852 (0.500–1.000) | 0.912 (0.899–0.926) |
| COAD | 86 | 0.947 (0.920–0.970) | 0.888 (0.874–0.903) | 0.907 (0.843–0.963) | 0.887 (0.872–0.902) |
| DLBC | 10 | 0.979 (0.951–0.996) | 0.875 (0.859–0.890) | 1 (1.000–1.000) | 0.874 (0.858–0.889) |
| ESCA | 36 | 0.957 (0.924–0.979) | 0.912 (0.898–0.925) | 0.944 (0.854–1.000) | 0.911 (0.897–0.924) |
| GBM | 6 | 0.965 (0.929–0.991) | 0.897 (0.882–0.911) | 1 (1.000–1.000) | 0.897 (0.882–0.911) |
| HNSC | 113 | 0.958 (0.928–0.980) | 0.931 (0.919–0.943) | 0.903 (0.843–0.955) | 0.933 (0.921–0.945) |
| KICH | 15 | 0.995 (0.989–1.000) | 0.97 (0.962–0.978) | 1 (1.000–1.000) | 0.97 (0.961–0.977) |
| KIRC | 84 | 0.984 (0.973–0.994) | 0.934 (0.922–0.945) | 0.952 (0.903–0.989) | 0.933 (0.921–0.945) |
| KIRP | 59 | 0.985 (0.973–0.994) | 0.908 (0.894–0.921) | 0.983 (0.943–1.000) | 0.905 (0.891–0.919) |
| LGG | 22 | 0.992 (0.979–0.999) | 0.984 (0.978–0.989) | 0.954 (0.850–1.000) | 0.984 (0.978–0.990) |
| LIHC | 81 | 0.972 (0.945–0.993) | 0.936 (0.924–0.947) | 0.951 (0.901–0.989) | 0.935 (0.923–0.947) |
| LUAD | 39 | 0.942 (0.912–0.968) | 0.851 (0.834–0.867) | 0.873 (0.750–0.972) | 0.85 (0.833–0.867) |
| LUSC | 33 | 0.931 (0.895–0.962) | 0.82 (0.801–0.838) | 0.909 (0.794–1.000) | 0.819 (0.800–0.836) |
| MESO | 11 | 0.958 (0.908–0.992) | 0.917 (0.904–0.929) | 0.91 (0.700–1.000) | 0.917 (0.904–0.930) |
| OV | 4 | 0.782 (0.632–0.985) | 0.637 (0.615–0.660) | 1 (1.000–1.000) | 0.636 (0.614–0.659) |
| PAAD | 49 | 0.978 (0.965–0.988) | 0.946 (0.935–0.956) | 0.918 (0.833–0.981) | 0.947 (0.936–0.957) |
| PCPG | 132 | 0.961 (0.942–0.977) | 0.935 (0.923–0.947) | 0.879 (0.821–0.933) | 0.94 (0.928–0.951) |
| PRAD | 97 | 0.989 (0.968–1.000) | 0.989 (0.984–0.994) | 0.99 (0.964–1.000) | 0.989 (0.984–0.994) |
| READ | 26 | 0.877 (0.778–0.955) | 0.894 (0.878–0.908) | 0.806 (0.643–0.952) | 0.895 (0.880–0.910) |
| SARC | 32 | 0.968 (0.947–0.986) | 0.836 (0.819–0.853) | 1 (1.000–1.000) | 0.833 (0.816–0.850) |
| SKCM | 43 | 0.952 (0.903–0.981) | 0.909 (0.896–0.923) | 0.906 (0.805–0.978) | 0.909 (0.896–0.923) |
| STAD | 112 | 0.891 (0.852–0.927) | 0.904 (0.889–0.917) | 0.777 (0.698–0.851) | 0.912 (0.898–0.926) |
| TGCT | 2 | 0.999 (0.995–1.000) | 0.997 (0.994–0.999) | 1 (1.000–1.000) | 0.997 (0.994–0.999) |
| THCA | 127 | 0.994 (0.988–0.998) | 0.972 (0.964–0.979) | 0.969 (0.934–0.993) | 0.972 (0.964–0.980) |
| THYM | 33 | 0.992 (0.986–0.997) | 0.931 (0.918–0.942) | 1 (1.000–1.000) | 0.929 (0.916–0.941) |
| UCEC | 91 | 0.978 (0.965–0.989) | 0.893 (0.879–0.907) | 0.966 (0.925–1.000) | 0.889 (0.874–0.904) |
| UCS | 8 | 0.871 (0.758–0.962) | 0.777 (0.758–0.797) | 0.871 (0.571–1.000) | 0.777 (0.757–0.797) |
| UVM | 24 | 0.999 (0.999–1.000) | 0.993 (0.989–0.996) | 1 (1.000–1.000) | 0.992 (0.988–0.996) |

### Survival analysis

Kaplan–Meier analysis of overall survival across all cancer types (external validation set) revealed a clear separation between the model-derived High– and Low-risk groups, with early divergence and a highly significant difference (p = 3.7 × 10⁻¹⁵). The High-risk group had a hazard ratio (HR) of 1.73 (95% CI: 1.51–1.98), indicating reasonable discrimination. Stratified Kaplan–Meier plots by cancer type (**Figure 3**) revealed consistent differences in survival rates and improved model performance for several individual cancers. The observed C-index values ranged from 0 to 0.8 across different cancer types, as depicted in **Supplementary Figure S6 (A)**.

**Figure 3.**
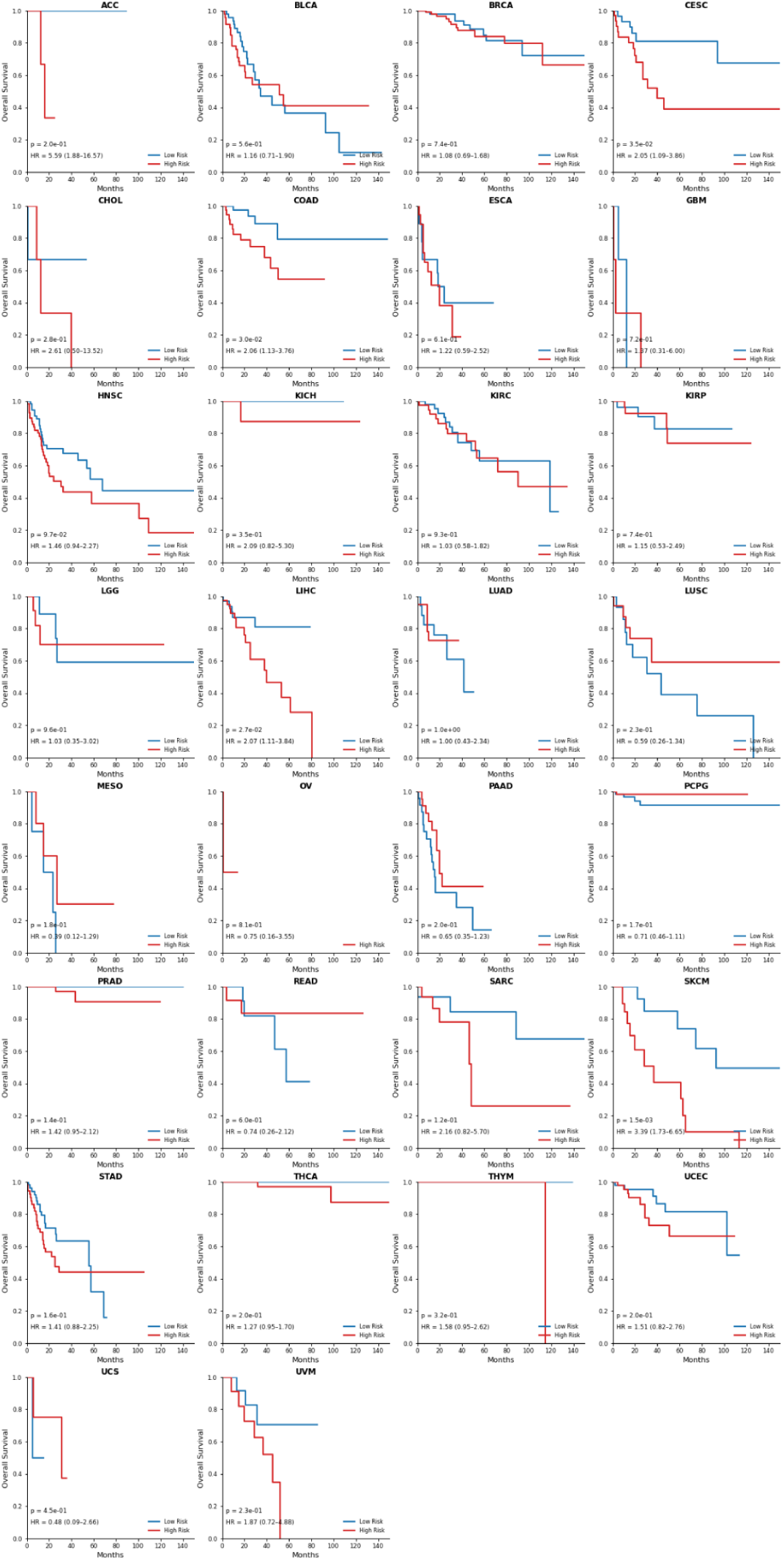
Kaplan–Meier survival analysis for various cancer types, showing survival curves for model-derived High– and Low-risk groups on the external validation set (1729 slides). Red and blue lines represent these risk groups over time (months).

**Figure 4.**
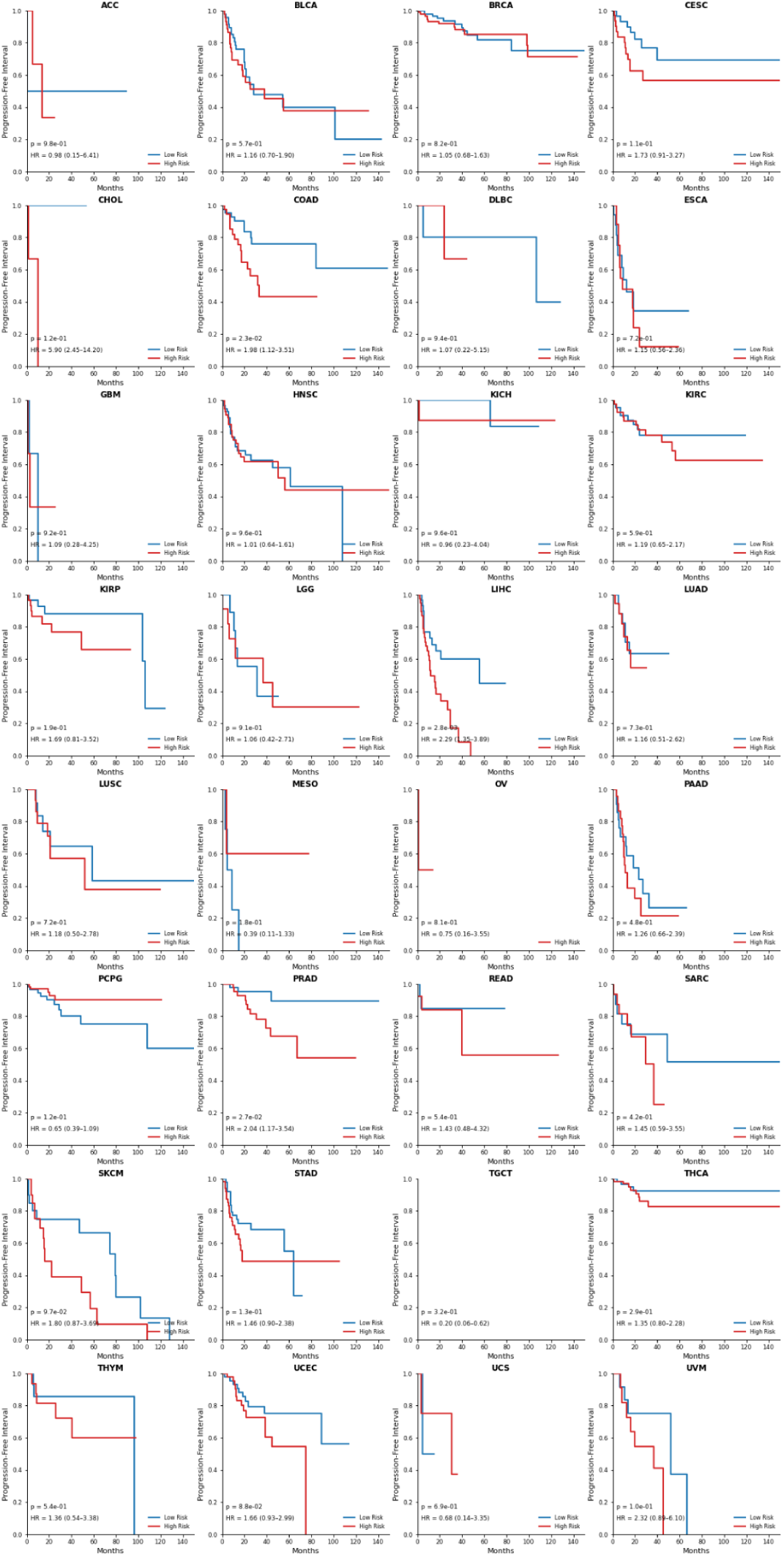
Kaplan-Meier analysis of Progression-Free Interval (PFI) comparing high-risk (red) and low-risk (blue) groups across different cancer types. Each plot shows the probability of being progression-free over time (months).

PFI analysis also showed overall strong separation between High– and Low-risk groups, with a significant difference confirmed by the log-rank test (p = 2.1 × 10⁻¹¹). The High-risk group had a hazard ratio of 1.56 (95% CI: 1.37–1.78), indicating a consistent association between predicted risk and disease progression. The C-index ranged from 0 to 0.8 (**Supplementary Figure S6 (B)**), indicating that most cancer classes exhibit distinguishable characteristics. The stratified Kaplan-Meier plots by cancer type (Figure 2) revealed clearer separation and improved model performance for almost all cancer types.

### Interpretability through attention and embeddings

In the external validation set, attention rollout from the model consistently focused on morphologically salient tumour regions. High-attention patches captured areas rich in tumour cells, nuclear atypia, and mitotic activity, while low-attention patches corresponded to stromal or sparsely cellular regions. Each panel presents slide-level predictions including cancer type, *TP53* mutation status with expression level, and OS/PFI outcomes, as illustrated in **Figure 5**. These results provide clinical context and confirm that predictions are based on histologically relevant features rather than global slide artifacts.

**Figure 5.**
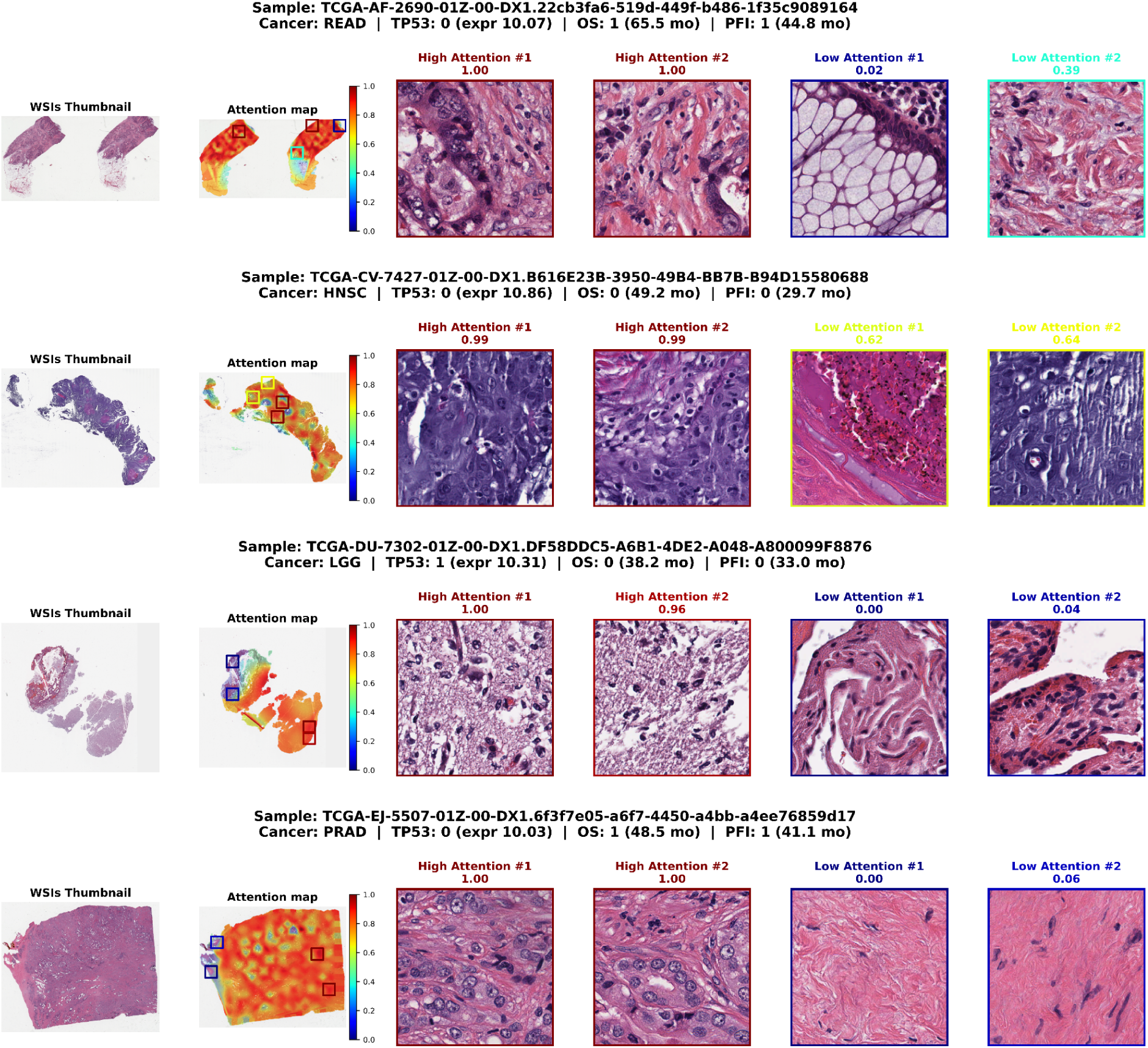
Slide-level attention across four Whole Slide Images (WSIs). Each row corresponds to one slide, showing the thumbnail, overlay attention, and the two highest-and two lowest-attention patches from the WSIs’ regions. Row headers summarise model predictions, including cancer type, TP53 mutation status, TP53 RNA expression levels, and clinical outcomes for overall survival (OS) and progression-free interval (PFI) events with time in months.

At the cohort level, global slide embeddings projected using t-distributed Stochastic Neighbor Embedding (t-SNE) revealed distinct, non-overlapping density clusters corresponding to predicted cancer types, while maintaining intra-cluster stratification based on *TP53* mutation status (*TP53* pathogenic variants versus wildtype or benign), as shown in **Figure 6**. Slides delineated with subtle black rings indicate low-margin predictions. These cases arise when the model’s top two cancer predictions exhibit comparable probabilities, which suggests a reduced level of classification confidence or the presence of overlapping morphological features. These visualisations offer complementary interpretability: patch-level attention highlights histologically salient regions, and cohort-level embedding structure reflects biologically coherent organisation of the model outputs. These visualisations underscore the alignment between learned representations and underlying pathology.

**Figure 6.**
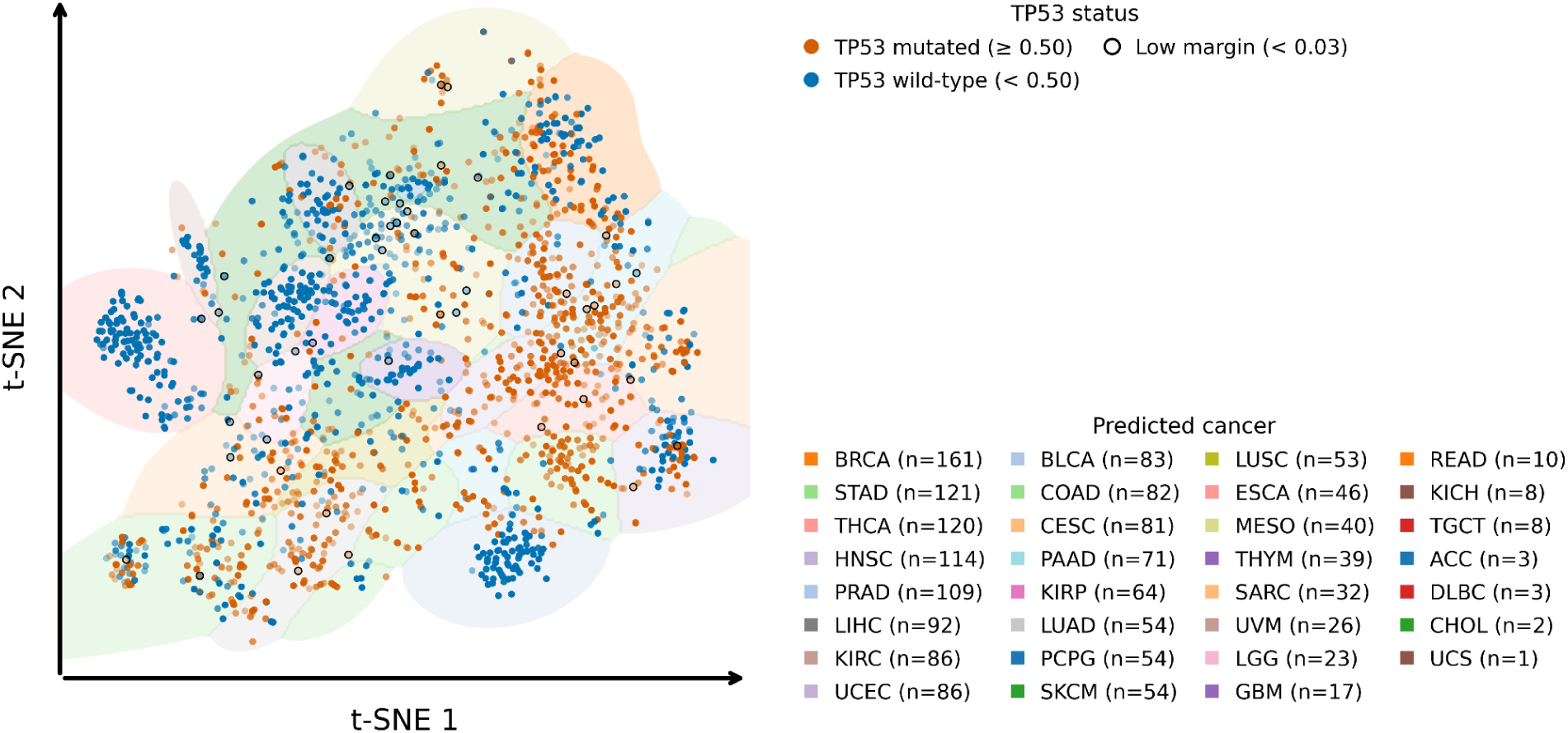
t-distributed Stochastic Neighbour Embedding (t-SNE) visualisation of slide-level feature embeddings from the multi-task fine-tuned model, with each point representing a WholeSlide Image (WSI) colored by TP53 mutation status (orange for mutated, blue for wild-type). The distinct clusters correspond to different cancer types, demonstrating morphological differences, while the mixed TP53 status within these clusters indicate shared histologic patterns across various cancers.

## DISCUSSION

Molecular profiling from routine histopathology could streamline precision oncology, yet integrating tumour classification, mutation status, gene expression, and prognosis remains challenging. Many existing models typically focus on a limited number of cancer types or use separate pipelines for these tasks, which restricts their clinical utility in oncology. In this study, we developed and evaluated a multi-task ViT-based MIL model for comprehensive pan-cancer analysis using histology images. Our model demonstrated robust performance across three key tasks: pan-cancer type classification, *TP53* biomarker detection, and survival prognostication. Our model demonstrates robust performance across diverse cancer types, using per-class Youden thresholds, with AUROC values exceeding 0.88 for all classes (**Table 2**) on the unseen WSIs (1729), except for ovarian tumours. These results indicate that a multi-resolution context fine-tuned model and refined thresholding are effective strategies for large-scale histopathology classification, supporting deployment across diverse tissue backgrounds and slide prediction.

In the molecular inference task, the model demonstrated reasonable performance for *TP53* mutation detection at the slide levels across 32 types of cancer, achieving an overall AUROC of 0.766 with a balanced accuracy of 0.72, sensitivity of 0.70, and specificity of 0.73. Our approach differs from prior mutation-focused frameworks, which combined self-supervised learning and attention MIL for pan-cancer mutation prediction across seven cancer types.^21^ While their model achieved an AUROC ≥ 0.70 for several genes and generalised well to external data, our multi-resolution ViT-based MIL framework extends beyond mutation inference to pan-cancer classification, prognostic risk stratification, and expression level estimation. Moreover, our interpretability strategy integrates patch-level attention with cohort-level embeddings (**Figures 5 and 6**), offering complementary insights into biological coherence. In prostate cancer, a deep-learning model for *TP53* achieved robust cross-cohort generalisation and provided spatial maps linking mutation status to aggressive phenotypes. In contrast, our approach prioritises mutation classification and embeds it within a multi-task framework, yielding a more comprehensive diagnostic tool.^45^

Hu et al. introduced CATfusion, a cross-attention transformer that integrates WSIs with multi-omics, achieving superior pan-cancer survival prediction (overall C-index ∼0.668) under five-fold cross-validation.^46^ Similarly, Wulczyn et al. demonstrated that deep learning can stratify survival risk across multiple cancer types, with incremental gains over baseline models.^47^ In contrast, our multi-task framework emphasises mutation classification (32 cancers; overall AUROC ∼0.77) and prognostic outcomes using only histopathology images, offering a deployable alternative when omics data are unavailable. Although our prognostic head identified survival-related patterns in histology, its reduced performance on external cohorts highlights the inherent limitations of relying solely on slides, likely due to missing clinical and molecular context. These findings highlight the need for multimodal integration and disease-specific architectures to achieve robust and generalisable survival prediction in real-world settings.

While this study demonstrates the potential of a ViT-based MIL framework for predicting molecular and clinical outcomes directly from WSIs, some limitations should be noted. First, the training datasets were highly imbalanced across tumour types, which may have affected the model’s ability to generalise, especially for underrepresented classes. Although class weighting and extensive data augmentation were applied to mitigate this issue, residual imbalance likely contributed to performance variability and reduced sensitivity in minority classes. Second, the model demonstrated limited prognostic performance, indicating that morphological features alone may not fully represent the complexity of survival-related factors. Next, the model was fine-tuned on downsampled images to reduce computational load and accelerate training, which may overlook finer-grained histological features or spatial context that could enhance the model’s performance. Furthermore, shared attention across multi-task heads may dilute task-specific focus, limiting the model’s ability to highlight regions relevant to *TP53* mutation. Finally, interpretability remains a challenge for the ViT-based MIL model in multi-task settings. While attention maps provide some insight into model focus, still required further validation to enhance clinical utility. Future work should explore integration with multi-modal data, broader external validation, and improved interpretability to enhance computational pathology.

In summary, the ViT-based multi-task MIL fine-tuned model demonstrated consistent generalisation to unseen WSIs, effectively capturing tumour type and mutation-related patterns, while showing limited predictive capacity for survival outcomes.

## DECLARATIONS SECTION

### ETHICAL APPROVAL AND CONSENT TO PARTICIPATE

The datasets used in this study were obtained from publicly accessible databases, thereby eliminating the need for direct patient interaction and the collection of personally identifiable information. Therefore, no additional ethical approval was required, and the study adheres to ethical guidelines and data-sharing policies.

## CONSENT FOR PUBLICATION

Not applicable

## AVAILABILITY OF SUPPORTING DATA

All the data used in this study are publicly accessible, and the links are provided.

## COMPETING INTERESTS

A.K.C., M.T.B., P.W.T., and A.W.H. are co-founders of Pandani Solutions Pty Ltd, which is developing tools for computational pathology.

## FUNDING

This work was supported by an Australian National Health and Medical Research Council Leadership Award (A.W.H.).

## AUTHORS’ CONTRIBUTIONS

A.K.C. designed the study, extracted and preprocessed data, selected the optimal architecture, implemented it for training and validating the models, analysed the results, and prepared the manuscript. P.W.T. and M.T.B. critically reviewed and interpreted the results. H.C.H. reviewed the results and provided interpretations within a pathological context. A.W.H. supervised the overall study design, reviewed the clinical significance and validation protocols, interpreted the analyses, provided valuable suggestions for improving the manuscript, and edited it. All authors reviewed the manuscript thoroughly and approved the final version.

## Data Availability

All the data used in this study are publically available.

https://portal.gdc.cancer.gov/

## ACKNOWLEDGEMENTS

We thank the University of Tasmania for providing the research infrastructure and resources that supported this study.

