## Supplemental information for "Prediction of *TP53* biomarkers and survival outcomes from whole slide images using a vision transformer-based multi-instance learning framework"

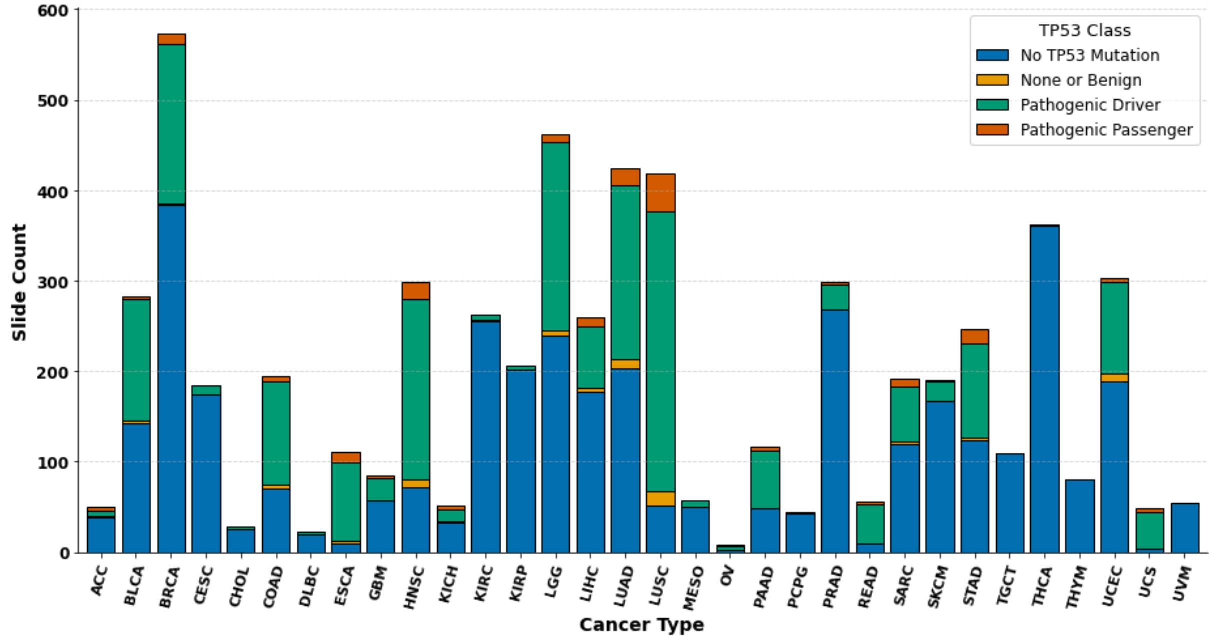


**Supplementary Figure S1** *Distribution of TP53 mutation status across 32 solid cancer types. The bar chart shows the number of whole-slide images (y-axis) for each cancer type (x-axis), stratified by TP53 mutation status: No TP53 Mutation (blue), None or Benign (yellow), Pathogenic Driver (green), and Pathogenic Passenger (orange). No mutation means the wildtype of TP53.*

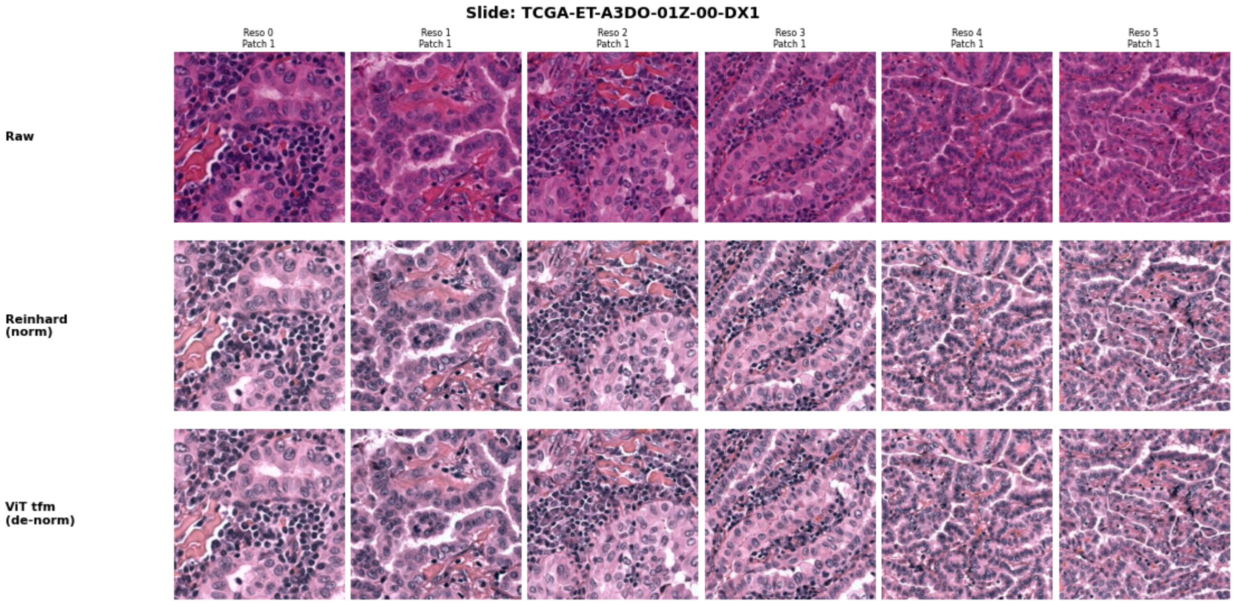


**Supplementary Figure S2** *The first row shows random patches selected from the TCGA-UT dataset, covering 32 tumour types at six magnifications (0.5–1.0 µm/pixel). The second row describes the Reinhard stain normalisation method, and the last row outlines the image transformations performed before model training.*

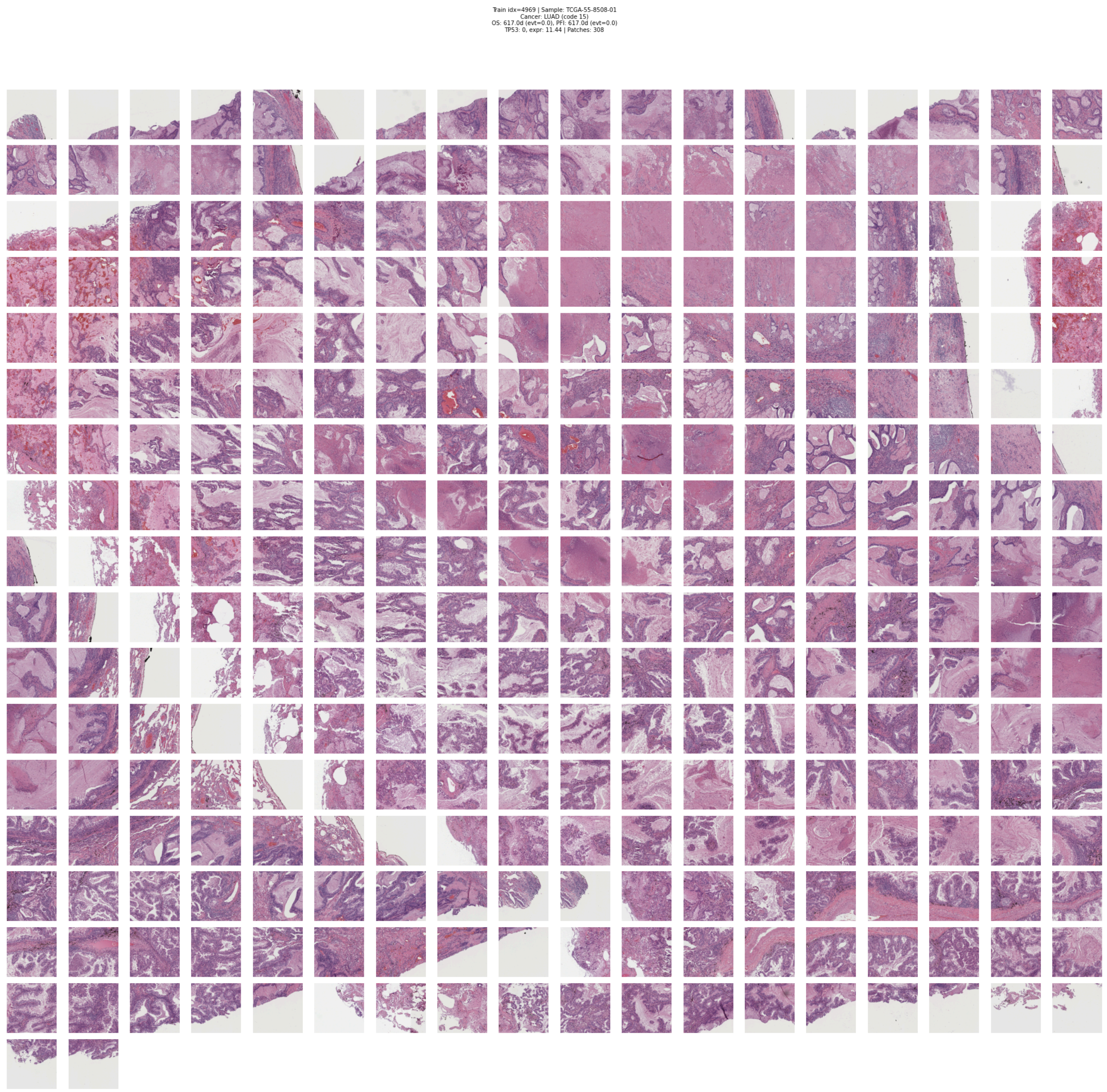


**Supplementary Figure S3** *illustrates all the extracted patches from a single whole-slide image (WSIs) at a resolution ( approximately 6x downsampled), including both cancerous and non-cancerous regions of WSIs. The cancer types, TP53 mutation status, TP53* RNA expression levels*, and survival outcomes are labelled (top) at the slide level.*

**Supplementary Table S1** *Multi-resolution ViT-based MIL model's performance on the validation set using Youden Index with class-specific thresholds.*

| Cancer Class | Slides number | AUROC | Accuracy | Sensitivity | Specificity |
|---|---|---|---|---|---|
| ACC | 10 | 0.972 (0.951–0.988) | 0.90 (0.883–0.917) | 1 (1.000–1.000) | 0.90 (0.883–0.916) |
| BLCA | 57 | 0.973 (0.957–0.986) | 0.922 (0.906–0.936) | 0.929 (0.855–0.983) | 0.921 (0.904–0.936) |
| BRCA | 115 | 0.961 (0.939–0.978) | 0.89 (0.872–0.908) | 0.938 (0.890–0.980) | 0.885 (0.866–0.904) |
| CESC | 37 | 0.939 (0.905–0.967) | 0.893 (0.876–0.910) | 0.84 (0.711–0.947) | 0.896 (0.877–0.914) |
| CHOL | 6 | 0.867 (0.737–0.991) | 0.654 (0.627–0.681) | 0.999 (1.000–1.000) | 0.653 (0.625–0.679) |
| COAD | 39 | 0.995 (0.991–0.998) | 0.97 (0.960–0.979) | 1 (1.000–1.000) | 0.968 (0.958–0.978) |
| DLBC | 4 | 0.995 (0.989–0.999) | 0.992 (0.986–0.997) | 0.98 (1.000–1.000) | 0.992 (0.986–0.997) |
| ESCA | 22 | 0.976 (0.959–0.990) | 0.923 (0.908–0.938) | 0.955 (0.850–1.000) | 0.923 (0.907–0.938) |
| GBM | 17 | 0.998 (0.996–1.000) | 0.991 (0.985–0.996) | 1 (1.000–1.000) | 0.991 (0.985–0.996) |
| HNSC | 60 | 0.986 (0.976–0.994) | 0.957 (0.946–0.968) | 0.933 (0.864–0.985) | 0.958 (0.947–0.970) |
| KICH | 9 | 0.983 (0.953–1.000) | 0.884 (0.866–0.901) | 1 (1.000–1.000) | 0.883 (0.864–0.900) |
| KIRC | 53 | 0.997 (0.993–1.000) | 0.978 (0.970–0.986) | 0.98 (0.933–1.000) | 0.978 (0.970–0.987) |
| KIRP | 41 | 0.99 (0.983–0.995) | 0.922 (0.907–0.936) | 1 (1.000–1.000) | 0.92 (0.904–0.936) |
| LGG | 92 | 0.999 (0.998–1.000) | 0.981 (0.974–0.988) | 0.989 (0.962–1.000) | 0.98 (0.972–0.988) |
| LIHC | 52 | 0.993 (0.986–0.998) | 0.987 (0.980–0.993) | 0.942 (0.875–1.000) | 0.989 (0.983–0.995) |
| LUAD | 84 | 0.98 (0.968–0.989) | 0.945 (0.932–0.957) | 0.905 (0.838–0.964) | 0.948 (0.934–0.961) |
| LUSC | 84 | 0.981 (0.965–0.991) | 0.94 (0.927–0.953) | 0.952 (0.904–0.989) | 0.94 (0.925–0.953) |
| MESO | 12 | 0.968 (0.940–0.992) | 0.874 (0.855–0.892) | 1 (1.000–1.000) | 0.872 (0.853–0.891) |
| OV | 1 | 0.883 (0.864–0.901) | 0.883 (0.865–0.900) | 0.627 (0.000–1.000) | 0.882 (0.864–0.901) |
| PAAD | 23 | 0.964 (0.912–0.998) | 0.985 (0.979–0.992) | 0.87 (0.714–1.000) | 0.987 (0.981–0.993) |
| PCPG | 9 | 0.993 (0.987–0.998) | 0.979 (0.970–0.986) | 1 (1.000–1.000) | 0.978 (0.970–0.986) |
| PRAD | 60 | 0.991 (0.981–0.998) | 0.964 (0.953–0.974) | 0.95 (0.889–1.000) | 0.965 (0.953–0.975) |
| READ | 11 | 0.974 (0.918–1.000) | 0.99 (0.984–0.995) | 0.913 (0.714–1.000) | 0.991 (0.985–0.996) |
| SARC | 38 | 0.997 (0.993–0.999) | 0.969 (0.959–0.979) | 1 (1.000–1.000) | 0.968 (0.957–0.978) |
| SKCM | 38 | 0.974 (0.936–0.993) | 0.966 (0.955–0.976) | 0.948 (0.868–1.000) | 0.967 (0.956–0.976) |
| STAD | 49 | 0.939 (0.906–0.967) | 0.944 (0.931–0.956) | 0.776 (0.652–0.900) | 0.951 (0.938–0.963) |
| TGCT | 20 | 0.989 (0.974–0.999) | 0.898 (0.882–0.914) | 1 (1.000–1.000) | 0.897 (0.880–0.914) |
| THCA | 72 | 0.998 (0.997–1.000) | 0.991 (0.985–0.996) | 0.973 (0.930–1.000) | 0.992 (0.986–0.997) |
| THYM | 15 | 0.999 (0.998–1.000) | 0.995 (0.991–0.998) | 1 (1.000–1.000) | 0.995 (0.991–0.998) |
| UCEC | 61 | 0.982 (0.973–0.989) | 0.889 (0.872–0.907) | 1 (1.000–1.000) | 0.883 (0.865–0.902) |
| UCS | 10 | 0.934 (0.855–0.990) | 0.944 (0.930–0.956) | 0.802 (0.500–1.000) | 0.945 (0.932–0.958) |
| UVM | 11 | 0.978 (0.954–0.996) | 0.89 (0.873–0.909) | 1 (1.000–1.000) | 0.889 (0.871–0.907) |

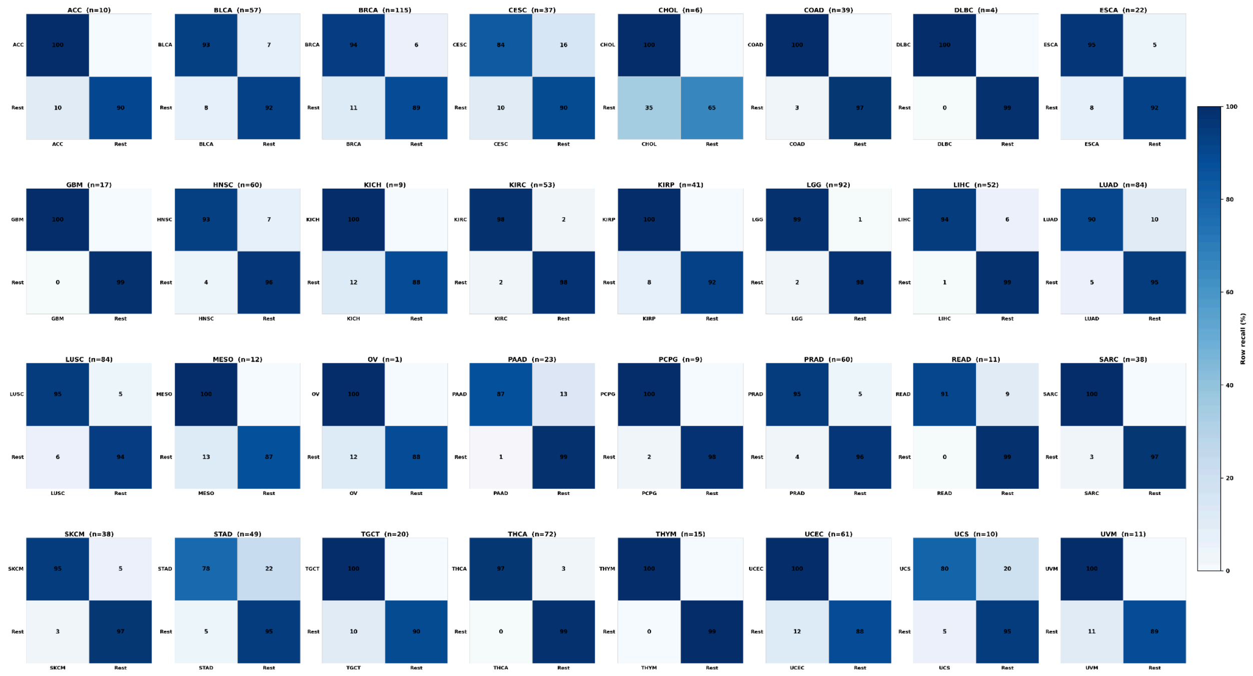

**Supplementary Figure S4** *One-versus-rest (OvR) classification performance across 32 solid tumour types, utilising the Multi-resolution ViT-based MIL model on the validation set.*

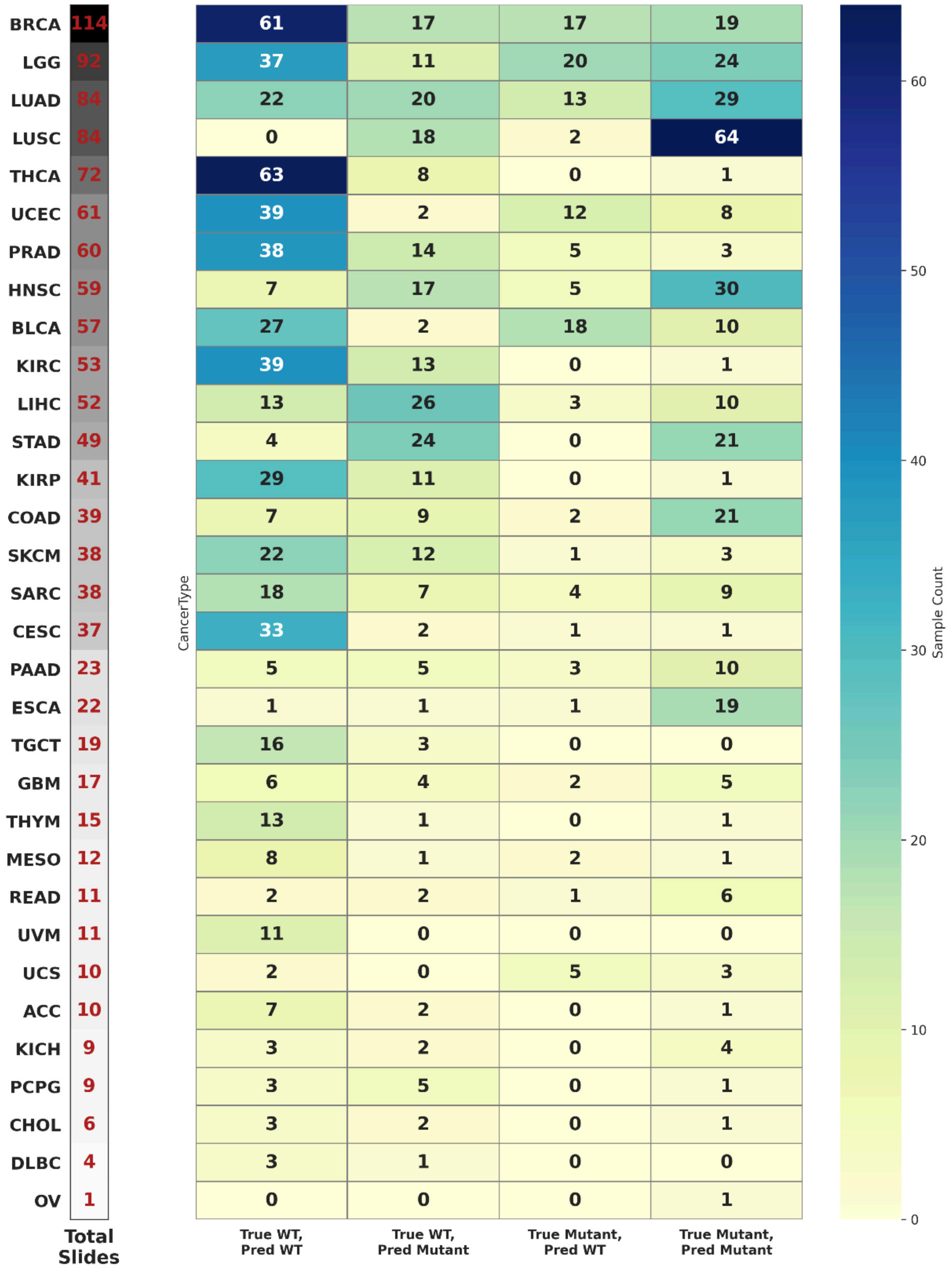


**Supplementary Figure S5** *Heatmap showing TP53 mutation prediction outcomes across cancer types. Each row represents a cancer type, annotated with its total number of Whole Slide Images (WSIs). Heatmap showing TP53 mutation prediction outcomes across cancer types, categorised by true and predicted wild-type or pathogenic variant (mutant) status.*

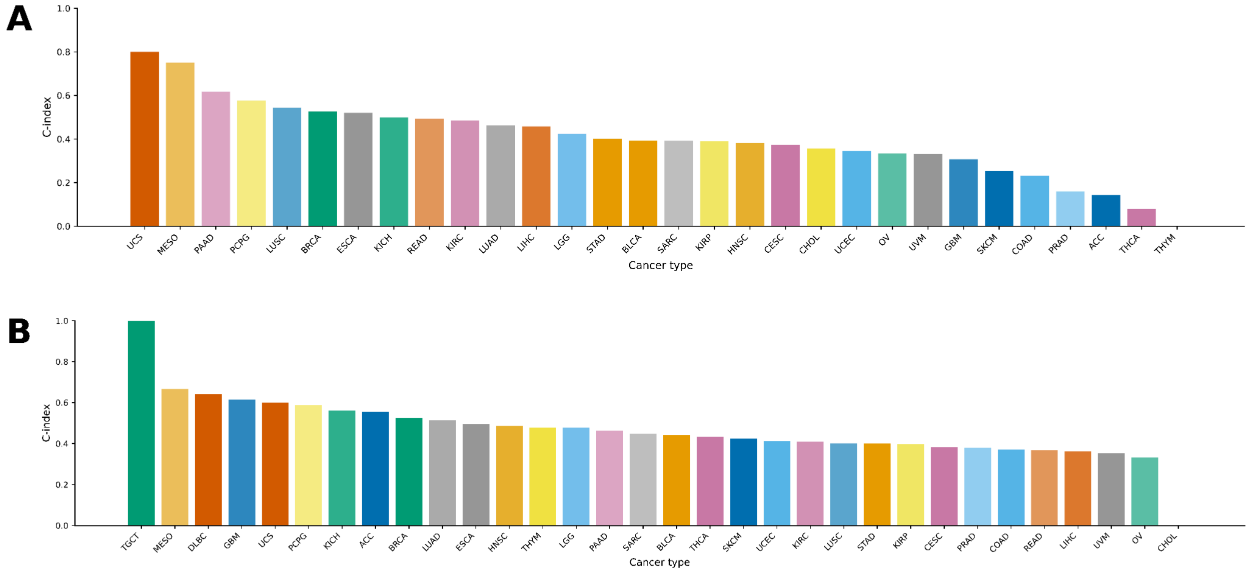


**Supplementary Figure S6** *Concordance performance by cancer type. (**A**) Overall Survival (OS) and (**B**) Progression-Free Interval (PFI) concordance index (C-index) for each tumour type. The Y-axis represents the C-index values, while the X-axis categorises different tumour types, highlighting differences in performance across cancers from the external validation set (1,729 slides) at the slide level*.
